# Obstructive Sleep Apnea is a Risk Factor for Incident COVID-19 Infection

**DOI:** 10.1101/2024.08.15.24312067

**Authors:** Stuart F. Quan, Matthew D. Weaver, Mark É. Czeisler, Laura K. Barger, Lauren A. Booker, Mark E. Howard, Melinda L. Jackson, Christine F. McDonald, Anna Ridgers, Rebecca Robbins, Prerna Varma, Shantha M.W. Rajaratnam, Charles A. Czeisler

**Affiliations:** Division of Sleep and Circadian Disorders, Brigham and Women’s Hospital, Boston, MA; Francis Weld Peabody Society, Harvard Medical School, Boston, MA; School of Psychological Sciences, Turner Institute for Brain and Mental Health, Monash University, Melbourne, Australia; Institute for Breathing and Sleep, Austin Health, Heidelberg, Victoria, Australia; University Department of Rural Health, La Trobe Rural Health School, La Trobe University, Bendigo, Victoria, Australia; Turner Institute for Brain and Mental Health, Monash University, Melbourne, Australia; Department of Medicine, The University of Melbourne, Melbourne, Victoria, Australia; Department of Respiratory and Sleep Medicine, Austin Health, Heidelberg, Victoria, Australia; Division of Sleep Medicine, Harvard Medical School, Boston, MA; Faculty of Medicine, Monash University, Melbourne Australia

**Keywords:** COVID-19, Obstructive Sleep Apnea, SARS-CoV-2 infection

## Abstract

Cross-sectional studies suggest that obstructive sleep apnea (OSA) is a potential risk factor for incident COVID-19 infection, but longitudinal studies are lacking. In this study, two surveys from a large general population cohort, the COVID-19 Outbreak Public Evaluation (COPE) Initiative, undertaken 147 ± 58 days apart were analyzed to determine whether the pre-existing OSA was a risk factor for the incidence of COVID-19. Of the 24,803 respondents completing the initial survey, 14,950 were negative for COVID-19; data from the follow-up survey were available for 2,325 respondents. Those with incident COVID-19 infection had a slightly higher prevalence of OSA (14.3 vs. 11.5%, p=0.068). Stratification by treatment status revealed that those untreated for their OSA were at greater risk for developing COVID-19 infection (OSA Untreated, 14.2 vs. 7.4%, p≤0.05). In a logistic regression model adjusted for comorbidities, demographic and socioeconomic factors and the interaction between vaccination status and OSA, incident COVID-19 infection was 2.15 times more likely in those with untreated OSA (aOR: 2.15, 95% CI: 1.18-3.92, p≤0.05). Stratification by treatment status revealed only untreated OSA participants were at greater risk for COVID-19 (aOR: 3.21, 95% CI: 1.25-8.23, p≤0.05). The evidence from this study confirms untreated OSA as a risk factor for acquiring COVID-19 infection and highlights the importance of actively treating and managing OSA as a preventative mechanism against COVID-19 disease.

## Introduction

After the onset of the COVID-19 pandemic, it rapidly became evident that infected individuals who were older and had medical comorbidities were at greater risk for more severe disease and higher mortality (Garg, 2020). Subsequently, studies identified obstructive sleep apnea (OSA) as one of the conditions associated with a greater rate of COVID-19 infection, hospitalization, and death (Hariyanto & Kurniawan, 2021). However, virtually all studies to date have been cross-sectional or retrospective and few have been conducted in population cohort studies. Whether OSA is associated with the development of incident COVID-19 infection is yet to be determined.

The purpose of this study was to determine in a general population cohort whether OSA is a risk factor for incident COVID-19 infection. To accomplish this, we used data from The COVID-19 Outbreak Public Evaluation (COPE) Initiative (http://www.thecopeinitiative.org/), a program focused on accumulating data on public attitudes about COVID-19 and its sequelae during the pandemic from large scale, demographically representative samples.

## Methods

### Study Design and Participants

From March 10, 2022 to October 15, 2022, the COPE Initiative administered five successive waves of surveys focused on accumulating data on the prevalence and sequelae of COVID-19 infection. Each wave consisted of approximately 5000 unique participants who were recruited using quota-based sampling to represent population estimates for age, sex, race, and ethnicity based on the 2020 U.S. census. Surveys were conducted online by Qualtrics, LLC (Provo, Utah, and Seattle, Washington, U.S. Informed consent was obtained electronically. From September 27, 2022 to December 31, 2022, all participants were recontacted and asked to complete a follow-up survey focused on obtaining information on COVID-19 subsequent to their initial survey. Responses to the follow-up survey were limited to approximately 5,000 individuals. The mean duration between surveys was 147 ± 58 days (Minimum 43, Maximum 284).

As shown in the Figure, of the 24,803 participants initially surveyed, 4,259 participated in the follow-up survey. Of these 4,259 respondents, 2,325 indicated on their initial survey that they neither had ever tested positive for COVID-19 infection nor had developed loss of smell or taste. Therefore, they constitute the analytic cohort for this report. The study was approved by the Monash University Human Research Ethics Committee (Study #24036).

### Survey Items

From the initial survey, participants self-reported their age, race, ethnicity, sex, height, weight, education level, employment status and household income. They also reported information on several current and past medical conditions by answering the question: “Have you ever been diagnosed with any of the following conditions?” In addition to OSA, opportunity was provided to endorse high blood pressure, cardiovascular disease (e.g., heart attack, stroke, angina), gastrointestinal disorder (e.g., acid reflux, ulcers, indigestion), cancer, chronic kidney disease, liver disease, sickle cell disease, chronic obstructive pulmonary disease and asthma. Possible responses to each condition were “Never”, “Yes I have in the past, but don’t have it now”, “Yes I have, but I do not regularly take medications or receiving treatment”, and “Yes I have, and I am regularly taking medications or receiving treatment”.

Symptoms of OSA were obtained from responses to the Pittsburgh Sleep Quality Index which was embedded into the initial survey and included items related to roommate or bedpartner reported “loud snoring” and “long pauses between breaths while you sleep” (Buysse et al., 1989). In addition, sleepiness was assessed from the following item in the questionnaire: During the past month, how often have you had trouble staying awake while driving, eating meals, or engaging in social activity”. Possible responses to all three items were “Not during the past month”, “less than once a week”, Once or twice a week” or “Three or more times a week”. Participants were considered to have symptoms of OSA at the time of their initial survey if they had either of the following combination of symptoms: 1) snoring “Three or more times a week”, and witnessed apnea or sleepiness “Once or twice a week”; 2) witnessed apnea and sleepiness “Once or twice a week”.

Ascertainment of past COVID-19 infection was obtained from both the initial and follow-up surveys using responses from the following questions related to COVID-19 testing or the presence of loss of taste or smell:

1. “Have you ever tested positive?”
2. “Have you experienced a problem with decreased sense of smell or taste at any point since January 2020?”

### Statistical Analyses

Results for continuous or ordinal variables are reported as their respective means and standard deviations (SD) and for categorical variables as their percentages. Participants were classified as having had a COVID-19 infection if they endorsed testing positive for COVID-19 or had experienced a loss of taste or smell. They were considered to have OSA if they endorsed currently having the condition whether treated or not, or if they had two or more symptoms of OSA. Vaccination status was dichotomized as Boosted (>2 vaccinations) or Not Boosted (≤2 vaccinations). Comorbid medical conditions were defined as currently having the condition whether treated or untreated. The effect of comorbid medical conditions was evaluated by summing the number of conditions reported by the participant (minimum value 0, maximum value 9). Body mass index (BMI) was calculated using self-reported height and weight as kg/m^2^. Socioeconomic covariates were dichotomized as follows: employment (retired vs. not retired), education (high school or less vs. at least some college) and income in U.S. Dollars (<$50,000 vs ≧$50,000).

After stratifying by COVID-19 infection status comparisons of continuous or ordinal variables were performed using Student’s unpaired t-test; categorical variables were compared using χ^2^. Multivariable logistic regression was utilized to determine whether OSA was associated with COVID-19 infection. Initially, a baseline model was constructed using only OSA. Subsequently, increasingly complex models were developed by sequentially including demographic factors, boosted vaccination status, boosted vaccination status and its interaction with OSA, comorbidities, and socioeconomic factors. An additional model stratified OSA into those who were treated and those who were not. All analyses were conducted using IBM SPSS version 29 (Armonk, NY).

## Results

In Table 1 is shown the association between incident COVID-19 infection status and obstructive sleep apnea, comorbid medical conditions, demographic, anthropometric and social characteristics. Of the 2,345 participants who were COVID-19 negative on the initial survey, 572 (55%) had incident COVID-19 infection (Figure). In comparison to those who remained COVID-19 infection negative at follow-up (N=1753, 45%), participants with incident COVID-19 infection were younger (60.5 ± 14.4 vs. 63.3 ± 13.0 years, p≤0.001) and had fewer medical comorbidities (1.2 ± 1.4 vs. 1.4 ± 1.4, p≤0.05). They also were less likely to be retired (50 vs. 56.4%, p≤.01) and to have a yearly income lower than the poverty level (23.8 vs. 29.0%, p≤0.01). Other characteristics, including percent receiving boosted vaccination and BMI, were not different between groups. Those with incident COVID-19 infection had a slightly higher prevalence of OSA (14.3 vs. 11.5%, p=0.068). When stratified by treatment status, those who were untreated for their OSA were at greater risk for developing COVID-19 infection (OSA Untreated, 14.2 vs. 7.4%, p≤0.05).

**Figure:**
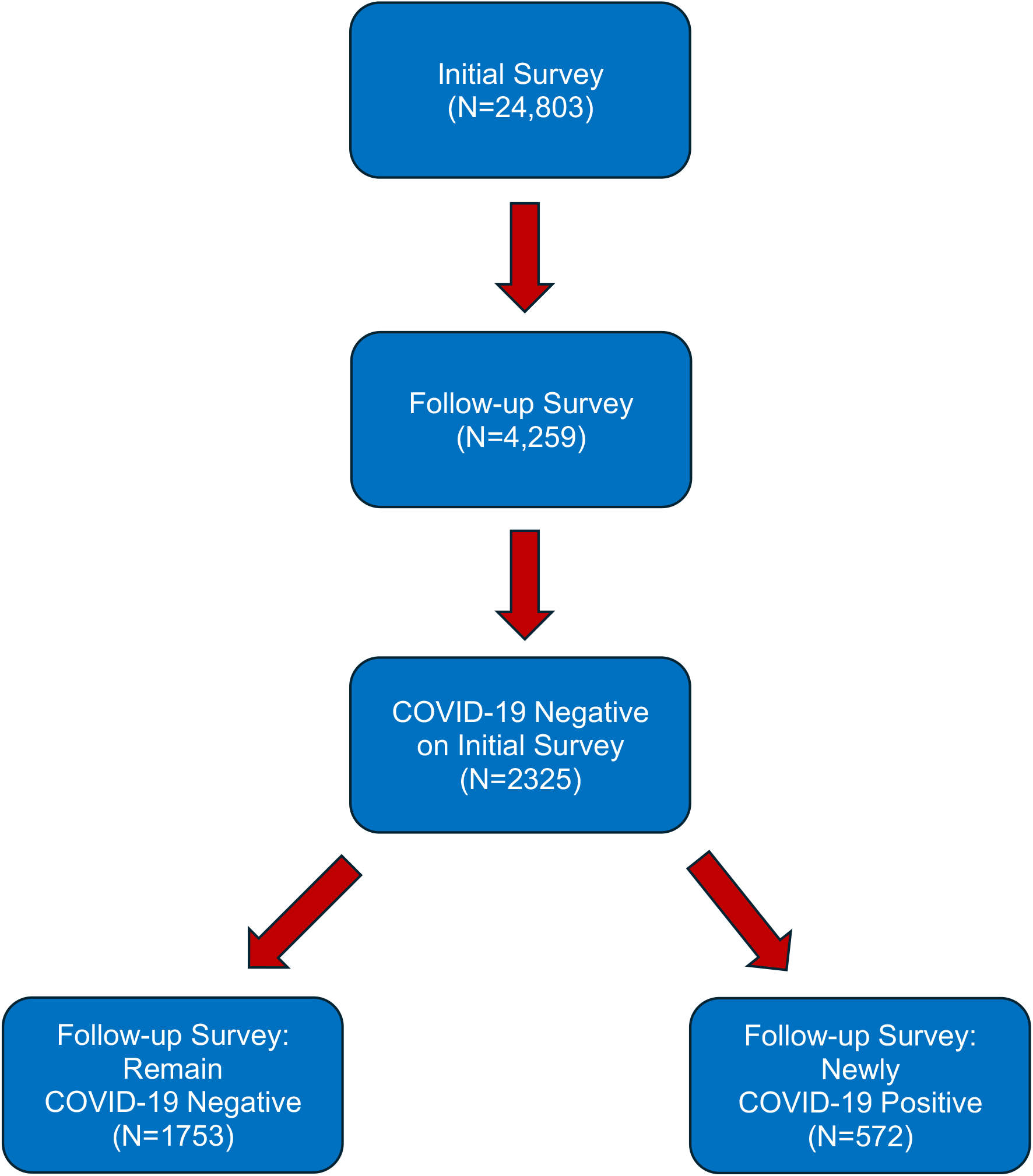
Participant Flow Chart

**Table 1:**
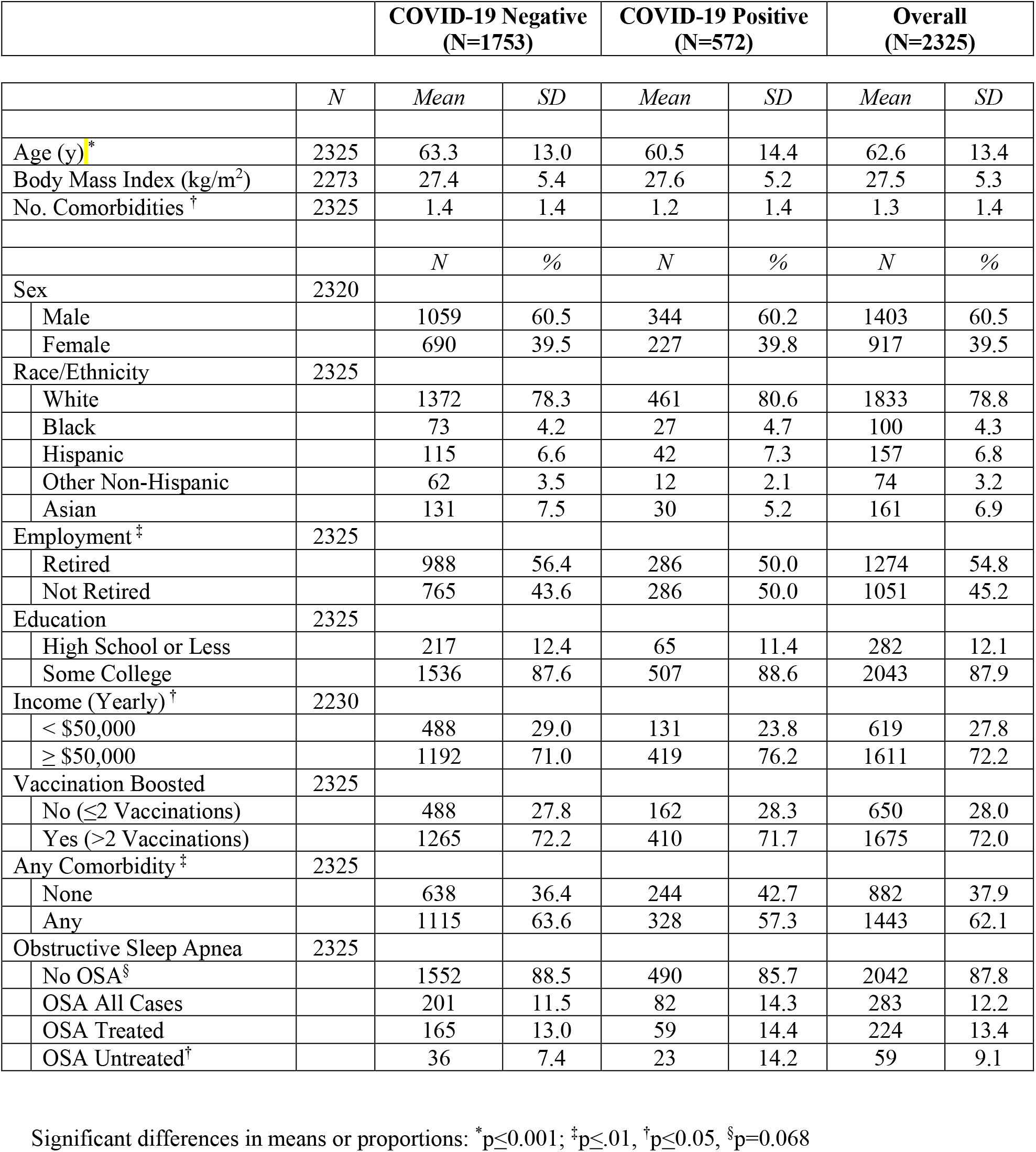
Associations Between COVID-19 Infection Status and Obstructive Sleep Apnea, Demographic, Social and Co-morbid Medical Characteristics.

Table 2 presents the logistic regression models for incident COVID-19 infection as a function of OSA as well as the interaction of OSA and boosted vaccination status. In the baseline model and in a model adjusted only for demographic factors, the aOR for OSA as a risk factor for incident COVID-19 infection approached statistical significance (aOR: 1.31, 95% CI: 0.99-1.73, p=0.07). However, after adjustment for the interaction of OSA and boosted vaccination status as well as for comorbidities and socioeconomic factors, those with OSA were 2.15 times more likely to develop COVID-19 infection (aOR: 2.15, 95% CI: 1.18-3.92, p≤0.05). Stratification by OSA treatment status demonstrated that only those with untreated OSA were at greater risk for COVID-19 infection (aOR: 3.21, 95% CI: 1.25-8.23, p≤0.05).

**Table 2:**
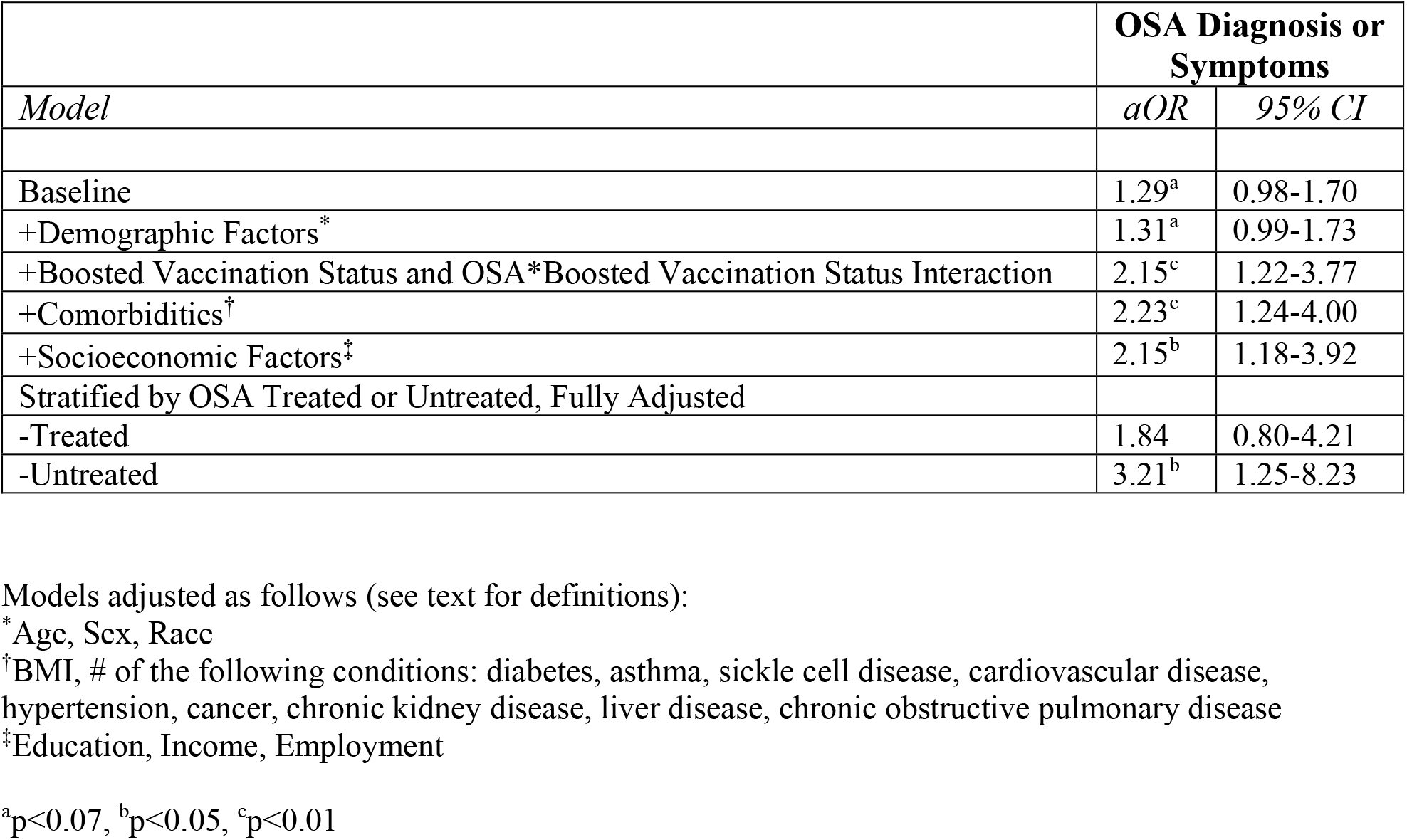
Adjusted Relative Odds of Incident COVID-19 Infection as a Function of Obstructive Sleep Apnea, Boosted Vaccination Status and their Interaction.

## Discussion

The principal finding of this longitudinal study is that untreated OSA is a risk factor for developing COVID-19 infection. Numerous previous cross-sectional investigations have observed a strong association between OSA and COVID-19 infection after adjusting for other co-morbid medical conditions including obesity (Garg, 2020). However, despite this extensive evidence associating OSA with COVID-19 infection, causation cannot be unambiguously assumed. Therefore, our finding extends these previous cross-sectional analyses by demonstrating that prospectively OSA is a risk factor for becoming infected with the SARS-CoV-19 virus.

In this study, stratification by treatment status, most likely continuous positive airway pressure (CPAP), indicated that participants who were untreated were at greater risk for COVID-19 infection related to OSA. This finding is consistent with a previous cross-sectional analysis of the COPE cohort (Quan et al., 2023), as well as other studies that also found that OSA treatment mitigated the impact of OSA on increasing risk of developing COVID-19 (Cade et al., 2020; Genzor et al., 2022). However, these observations are in contradistinction to a recent retrospective analysis of a large healthcare administrative data base in which OSA patients who had been prescribed continuous positive airway pressure developed COVID-19 infection at a greater rate than those who presumptively did not have OSA (Kendzerska et al., 2023). Prior treatment with CPAP also did not mitigate adverse outcomes in patients hospitalized with COVID-19 (Sampol et al., 2022). Thus, it is unclear whether treated individuals with OSA are protected; further studies are indicated to address this issue.

Several mechanisms may explain the increase in risk for developing COVID-19 conferred by OSA. Intermittent hypoxemia occurring during apneic or hypopneic events can promote the release of inflammatory cytokines (Ren & Hu, 2017). Intermittent hypoxemia also produces oxidative stress with release of reactive oxygen species further contributing to inflammation (Stanek et al., 2021). Introduction of the SARS-CoV-2 virus into a pre-existing inflammatory milieu could increase the likelihood of becoming infected. In addition, OSA also has been associated with cellular immune dysfunction and therefore may be a contributing factor to greater susceptibility to COVID-19 infection (Ludwig et al., 2022).

There are important limitations to this study. First, ascertainment of COVID-19 infection, OSA, and the presence of covariates were by self-report which may have resulted in misclassification. Second, only 17% of our initial participants were included in our follow-up survey which increases the likelihood that some selection bias occurred. Our finding that COVID-19 positive participants were less likely to have a comorbid condition suggests that our sample may not be representative of the entire COPE cohort and by extension, the general adult U.S. population. Nevertheless, despite a healthier cohort, we found that OSA was a risk factor for COVID-19 infection.

In conclusion, using a longitudinal cohort, OSA is a risk factor for the development of COVID-19 infection, thus confirming findings from previous cross-sectional studies. Treatment of OSA mitigated this risk, highlighting its importance in potentially preventing COVID-19. Additional studies using larger cohorts with objective ascertainment of COVID-19 infection should be performed.

## Data Availability

All data produced in the present study are available upon reasonable request to the authors

## Acknowledgments

Concept and Design: SFQ

Data collection: MDW, MÉC, MEH

Data analysis and interpretation: SFQ, MDW

Drafting of the manuscript: SFQ

Critical feedback and revision of manuscript: SFQ, MDW, MÉC, LKB, LAB, MEH, MLJ, CFM, AR, RR, PV, SMWR, CAC

## References

Buysse, D. J., Reynolds, C. F., Monk, T. H., Berman, S. R., & Kupfer, D. J. (1989). The Pittsburgh Sleep Quality Index: A new instrument for psychiatric practice and research. Psychiatry Research, 28(2), 193–213. 10.1016/0165-1781(89)90047-4

Cade, B. E., Dashti, H. S., Hassan, S. M., Redline, S., & Karlson, E. W. (2020). Sleep Apnea and COVID-19 Mortality and Hospitalization. American Journal of Respiratory and Critical Care Medicine, 202(10), 1462–1464. 10.1164/rccm.202006-2252LE

Garg, S. (2020). Hospitalization Rates and Characteristics of Patients Hospitalized with Laboratory-Confirmed Coronavirus Disease 2019—COVID-NET, 14 States, March 1– 30, 2020. MMWR. Morbidity and Mortality Weekly Report, 69(15), 458–464. 10.15585/mmwr.mm6915e3

Genzor, S., Prasko, J., Mizera, J., Jakubec, P., Sova, M., Vanek, J., Šurinová, N., & Langova, K. (2022). Risk of Severe COVID-19 in Non-Adherent OSA Patients. Patient Preference and Adherence, 16, 3069–3079. 10.2147/PPA.S387657

Hariyanto, T. I., & Kurniawan, A. (2021). Obstructive sleep apnea (OSA) and outcomes from coronavirus disease 2019 (COVID-19) pneumonia: A systematic review and meta-analysis. Sleep Medicine, 82, 47–53. 10.1016/j.sleep.2021.03.029

Kendzerska, T., Povitz, M., Gershon, A. S., Ryan, C. M., Talarico, R., Franco Avecilla, D. A., Robillard, R., Ayas, N. T., & Pendharkar, S. R. (2023). Association of clinically significant obstructive sleep apnoea with risks of contracting COVID-19 and serious COVID-19 complications: A retrospective population-based study of health administrative data. Thorax, 78(9), 933–941. 10.1136/thorax-2022-219574

Ludwig, K., Huppertz, T., Radsak, M., & Gouveris, H. (2022). Cellular Immune Dysfunction in Obstructive Sleep Apnea. Frontiers in Surgery, 9, 890377. 10.3389/fsurg.2022.890377

Quan, S. F., Weaver, M. D., Czeisler M.É., Barger, L. K., Booker, L. A., Howard, M. E., Jackson, M. L., Lane, R., McDonald, C. F., Ridgers, A., Robbins, R., Varma, P., Rajaratnam, S. M. W., & Czeisler, C. A. (2023). Associations between obstructive sleep apnea and COVID-19 infection and hospitalization among US adults. Journal of Clinical Sleep Medicine: JCSM: Official Publication of the American Academy of Sleep Medicine, 19(7), 1303–1311. 10.5664/jcsm.10588

Ren, H., & Hu, K. (2017). Inflammatory and oxidative stress-associated factors in chronic intermittent hypoxia in Chinese patients, rats, lymphocytes and endotheliocytes. Molecular Medicine Reports, 16(6), 8092–8102. 10.3892/mmr.2017.7632

Sampol, J., Sáez, M., Martí, S., Pallero, M., Barrecheguren, M., Ferrer, J., Sampol, G., & Vall d’Hebron COVID-19 Working Group. (2022). Impact of home CPAP-treated obstructive sleep apnea on COVID-19 outcomes in hospitalized patients. Journal of Clinical Sleep Medicine: JCSM: Official Publication of the American Academy of Sleep Medicine, 18(7), 1857–1864. 10.5664/jcsm.10016

Stanek, A., Brożyna-Tkaczyk, K., & Myslinski, W. (2021). Oxidative Stress Markers among Obstructive Sleep Apnea Patients. Oxidative Medicine and Cellular Longevity, 2021, 9681595. 10.1155/2021/9681595

